# Structural Signatures of Gender Norms: Cross-National Predictability of Attitudes Justifying Violence Against Women

**DOI:** 10.64898/2026.01.25.26344795

**Authors:** Caroline L. Alves

## Abstract

Violence against women is sustained not only by individual behavior but also by social norms that legitimize coercion and control. While attitudes justifying intimate-partner violence have been extensively documented in large-scale household surveys, they are rarely analyzed as structured, predictable population-level phenomena. Here, we model the continuous prevalence of violence-justifying attitudes across 70 countries and demographic subgroups using country-resolved supervised machine learning with strict out-of-sample evaluation. Drawing on harmonized estimates derived from the Demographic and Health Surveys, we quantify how much cross-subgroup variation in normative acceptance is explainable from survey structure alone. By comparing full models that incorporate attitudinal scenario framing with demographics-only baselines, we show that high predictability arises from fundamentally different sources across countries: in some contexts, demographic stratification—particularly education—structures normative acceptance, whereas in others, conditional justification narratives dominate. Integrating independent country-level indicators of gender inequality, human development, and democratic quality reveals that violence-justifying norms are most predictable in structurally polarized settings rather than within a single cultural regime. Together, these findings demonstrate that normative acceptance of violence is not uniformly diffuse but can form coherent, structurally embedded patterns. This cross-scale framework provides a quantitative basis for identifying where prevention strategies may benefit most from demographic targeting versus direct challenges to context-specific justifications of violence.

**Significance statement:** Normative acceptance of intimate-partner violence is a measurable societal risk factor, yet it is rarely analyzed as a structured population-level phenomenon. Most quantitative studies remain descriptive, and machinelearning analyses using large-scale household surveys typically focus on individual-level classification of victimization or vulnerability. Here, we model the continuous prevalence of violence-justifying attitudes across 70 countries and demographic subgroups using country-resolved supervised regression with rigorous out-of-sample evaluation. By contrasting demographics-only models with those incorporating attitudinal scenario framing, we show that cross-national differences in predictability arise from distinct sources—demographic stratification in some contexts and conditional justification narratives in others. Linking these patterns to independent indicators of gender inequality, human development, and democratic quality reveals that highly structured norms emerge in structurally polarized settings, highlighting where targeted prevention strategies are most likely to be effective.

## 1 Introduction

Violence against women remains a pervasive human rights violation and a major public health concern worldwide [22, 21]. The World Health Organization and the United Nations indicate that approximately 30% of women globally, around 840 million individuals, have experienced physical or sexual violence during their life-time [69]. Furthermore, data from 2023–2024 suggest that more than 300 million women were affected within a single year, pointing to limited progress in prevention [70]. Beyond non-lethal outcomes, femicide, which is the most extreme manifestation of gender-based violence, claimed roughly 50 000 lives worldwide in 2024, with nearly 60% of these perpetrated by intimate partners or family members [62]. Together, these patterns reflect durable structural gender inequalities and social norms that continue to legitimize violence within private and domestic spheres [29, 36].

To better understand this problem, existing quantitative research into the attitudinal justification of intimate partner violence has historically relied on large-scale household surveys, most notably the Demographic and Health Surveys (DHS) [14, 38]. This literature has remained almost exclusively inferential, focusing on the socio-demographic correlates of violence acceptance and their relative association strengths [61, 16, 64, 55]. While seminal multi-country analyses have identified education, employment, and household structure as key drivers of this heterogeneity across low- and middle-income contexts [64, 61], the analytical scope has been constrained. Even sophisticated multi-level and socio-ecological frameworks have prioritized parameter-driven inference—specifically the estimation of Odds Ratios (OR) for demographic correlates such as maternal education or rural residence—over out-of-sample predictive utility [63, 17]. While these models effectively identify statistically significant associations, they lack the algorithmic robustness to forecast whether an individual or a new sub-population will endorse violence, a gap that limits their efficacy for targeted behavioral interventions. Consequently, while reviews confirm that structural inequalities and social norms underpin these attitudes, the field lacks formal predictive benchmarks or rigorous model comparisons, leaving a critical gap in our ability to forecast attitudinal shifts [66].

More recently, a complementary line of work has applied machine learning (ML) methods to DHS- and NFHS-type survey microdata, where NFHS refers to the National Family Health Survey, the Indian implementation of DHS. These studies typically reframe gender-based violence as an individual-level classification problem—for example, predicting experience of intimate partner violence (IPV), vulnerability status, or related binary outcomes—and evaluate performance using metrics such as sensitivity, accuracy, or area under the receiver operating characteristic curve (AUC). While achieving high accuracy and performance, this classification-centric approach prioritizes binary threshold discrimination over the Coefficient of Determination (*R*^2^). By focusing on classification accuracy rather than *R*^2^, these studies effectively obscure the total proportion of societal-level attitudinal variation that can be systematically attributed to underlying survey structure. For instance, Shashidhara et al. used India’s NFHS-4 (2015–2016) to develop an ML-based IPV screening tool that compresses a high-dimensional questionnaire into a short instrument, reporting a recall of approximately 0.78 for identifying high-risk women [56]. Rahman et al. benchmarked multiple classifiers on the 2019–2020 Liberia DHS and reported best-case accuracies around 0.82 for predicting domestic-violence vulnerability, highlighting the effectiveness of ensemble methods such as LightGBM and Random Forest for tabular survey data [51]. Related ML studies using NFHS/DHS-style inputs— predicted across countries and demographics [52], ML-based hypothesis generation for child marriage [53], and supervised modeling of downstream behavioral responses such as help-seeking [15]—similarly emphasize individual-level risk stratification and variable-importance discovery.

In contrast to prior machine-learning studies that typically frame gender-based violence as an individual-level classification problem, the present study addresses a distinct but complementary question: how well can the population-level prevalence of violence-justifying attitudes be predicted across countries and demographic subgroups? Rather than modeling individual victimization or vulnerability, we treat normative acceptance of intimate-partner violence as a continuous societal quantity and apply country-resolved supervised regression to quantify the proportion of its variation that is explainable from survey structure alone (Materials and Methods; Section 2). The dataset analyzed here is a harmonized compilation derived from the DHS Program, summarizing country–year–subgroup prevalence of violence justification under specific scenarios (Section 2.1.1). This approach addresses two persistent gaps. First, existing ML work rarely evaluates the out-of-sample predictability of attitudinal endorsement at the country level, despite its central role in shaping behavioral norms and policy environments. Second, micro-level predictive structure is seldom linked to macro-structural context using harmonized external indicators. To address these limitations, we implement country-specific regression models under two specifications: a full model incorporating demographic composition, attitudinal framing, and time (Part I; Section 3.1), and a demographics-only baseline excluding attitudinal items (Part II; Section 3.2). We assess generalization using held-out *R*^2^ and error metrics and interpret model behavior via feature-family attribution. Finally, we integrate independent country-level indicators spanning cultural values, gender inequality, human development, and institutional quality, including Hofstede’s cultural dimensions [30], the United Nations Development Programme Human Development Reports [3], and the Varieties of Democracy (V-Dem) project [13] as disseminated via Our World in Data (Part III; Sections 2.1.2 and 3.3)—to test whether high predictability concentrates within shared macro-structural regimes, thereby establishing a cross-scale link between normative endorsement patterns and structural inequality. Table 1 summarizes prior quantitative studies alongside the present work, highlighting differences in data, analytical scope, modeling strategy, and evaluation criteria.

**Table 1:** Prior quantitative and machine-learning studies on gender-based violence using DHS/NFHS-style data, contrasted with the present work. Earlier DHS-based studies predominantly adopt *inferential regression* frameworks to estimate associations between socio-demographic factors and attitudes toward intimate-partner violence, without out-of-sample predictive evaluation. More recent machine-learning (ML) approaches typically reframe gender-based violence as an *individual-level classification* problem (e.g., victimization, vulnerability, or help-seeking) and assess performance using accuracy-, sensitivity-, or AUC-based metrics. In contrast, the present study models the *population-level prevalence of violence-justifying attitudes* via country-resolved supervised regression, evaluated with held-out *R*^2^ and error metrics and contextualized using macro-structural indicators. **Abbreviations:** DHS, Demographic and Health Surveys; NFHS, National Family Health Survey (India); IPV, intimate partner violence; ML, machine learning; RF, Random Forest; GBM, Gradient Boosting Machine; ANN, artificial neural network; DT, decision tree; AUC, area under the receiver operating characteristic curve; *R*^2^, coefficient of determination; MAE, mean absolute error; RMSE, root mean squared error.

| Study | Data and outcome | Modeling paradigm and evaluation |
| --- | --- | --- |
| Uthman et al. (2009) | DHS; justification of wife beating (17 sub-Saharan African countries) | Inferential multivariable regression; odds ratios and confidence intervals; no predictive validation |
| Tran et al. (2016) | DHS; acceptance of IPV (39 low- and middle-income countries) | Cross-national inferential regression; descriptive and coefficient-based interpretation |
| Doku & Asante (2015) | DHS; approval of domestic physical violence (Ghana) | Single-country logistic regression; odds ratios; no predictive or out-of-sample evaluation |
| Shashidhara et al. (2024) | NFHS-4 (India); individual IPV experience (binary) | ML classification (RF, GBM, logistic regression); sensitivity/recall $\approx 0.78$; questionnaire dimensionality reduction |
| Rahman et al. (2023) | Liberia DHS; individual domestic-violence vulnerability (binary) | ML classification (ANN, RF, DT, XGBoost, LightGBM); accuracy $\approx 0.82$ |
| McDougal et al. (2021) | NFHS-4 (India); non-marital sexual violence (binary) | Regularized ML and neural networks; individual risk stratification; no country-level prediction |
| Raj et al. (2020, 2021) | NFHS-4 (India); child marriage and related harmful practices | ML-based hypothesis generation; variable-importance discovery; classification framework |
| Dehingia et al. (2022) | DHS/NFHS; help-seeking following domestic violence (binary) | Supervised ML classification; accuracy- and AUC-based evaluation |
| **This study** | DHS-derived harmonized country-year-subgroup *prevalence* of violence-justifying attitudes (continuous; 70 countries) | Country-resolved supervised regression; held-out $R^2$, and other regression metrics; feature-family attribution; integration with macro-structural indicators |

**Table 2:**
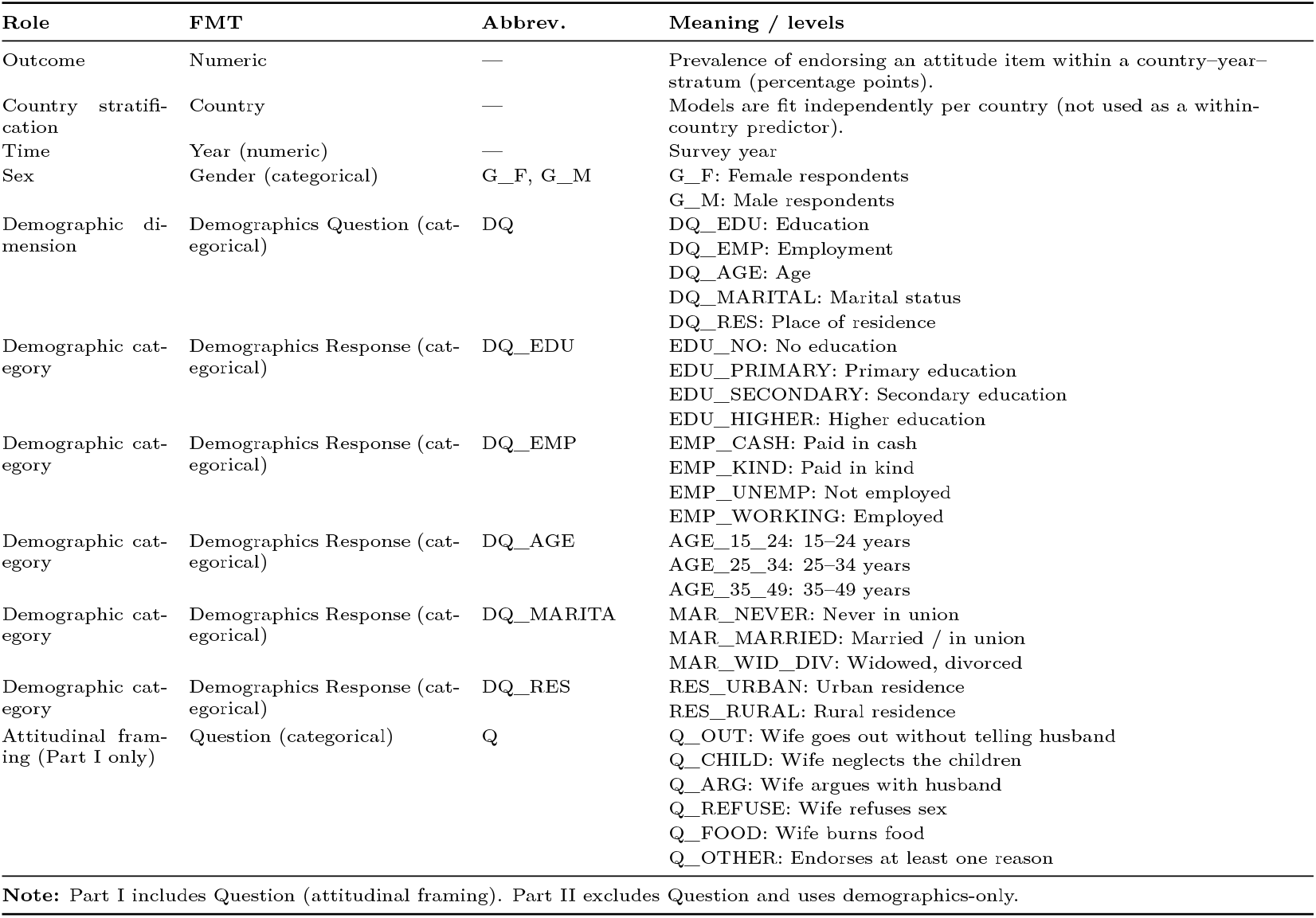
Variables used in Parts I–II (cleaned modeling table), abbreviation codes used in figures, and their meanings. **Abbreviations:** FMT, Field in modeling table; Abbrev., Abbreviation.

Against this background, the present study addresses three interrelated questions:

- **Predictability of norms**: How predictable is the population-level prevalence of violence-justifying attitudes across countries and demographic sub-groups when modeled as a continuous societal quantity?
- **Sources of explanatory structure**: How much predictive information is contributed by demographic composition alone, and how much is added by attitudinal framing of violence-related scenarios?
- **Cross-scale embedding**: Do countries in which violence-justifying attitudes are highly predictable occupy shared macro-structural contexts—spanning gender inequality, human development, democratic quality, and cultural values—or are they distributed across distinct structural regimes?

## 2 Materials and methods

### 2.1 Data

#### 2.1.1 Survey dataset and analytical decomposition

We analyzed a cleaned modeling table derived from a public harmonized compilation of cross-national survey estimates on attitudes justifying intimate-partner violence against women.^1^ The compilation aggregates large-scale household survey responses into country-level prevalence estimates for a standard set of attitude items commonly operationalized as whether a husband is justified in hitting or beating his wife if she (i) goes out without telling him, (ii) neglects the children, (iii) argues with him, (iv) refuses to have sex with him, or (v) burns the food, as well as a composite indicator of endorsing violence for at least one specified reason. Each record in the cleaned table corresponds to one country–year–stratum–item prevalence value (in percentage points).

The cleaned modeling table contains 11,187 records spanning 70 countries, with one survey year per country (years 2000–2018). Predictors comprise respondent sex (female, male), one demographic stratification dimension (demographics question: education, employment, age, marital status, or residence), one corresponding category within that dimension (demographics response), the attitude item (question), and survey year. The outcome is the estimated prevalence of endorsing the attitude item within the specified stratum, expressed in percentage points.

The harmonized design yields up to 2 × 15 × 6 = 180 records per country (two genders, 15 demographic-response strata, six attitude items). After validity filtering in the preprocessing pipeline (consistent country identifiers; non-missing predictors and outcome; consistent year parsing), per-country sample sizes range from 78 to 180 (median = 180; interquartile range = 163–180), reflecting heterogeneous completeness across country surveys. Figure 1 reports country-level record counts after cleaning.

**Figure 1:**
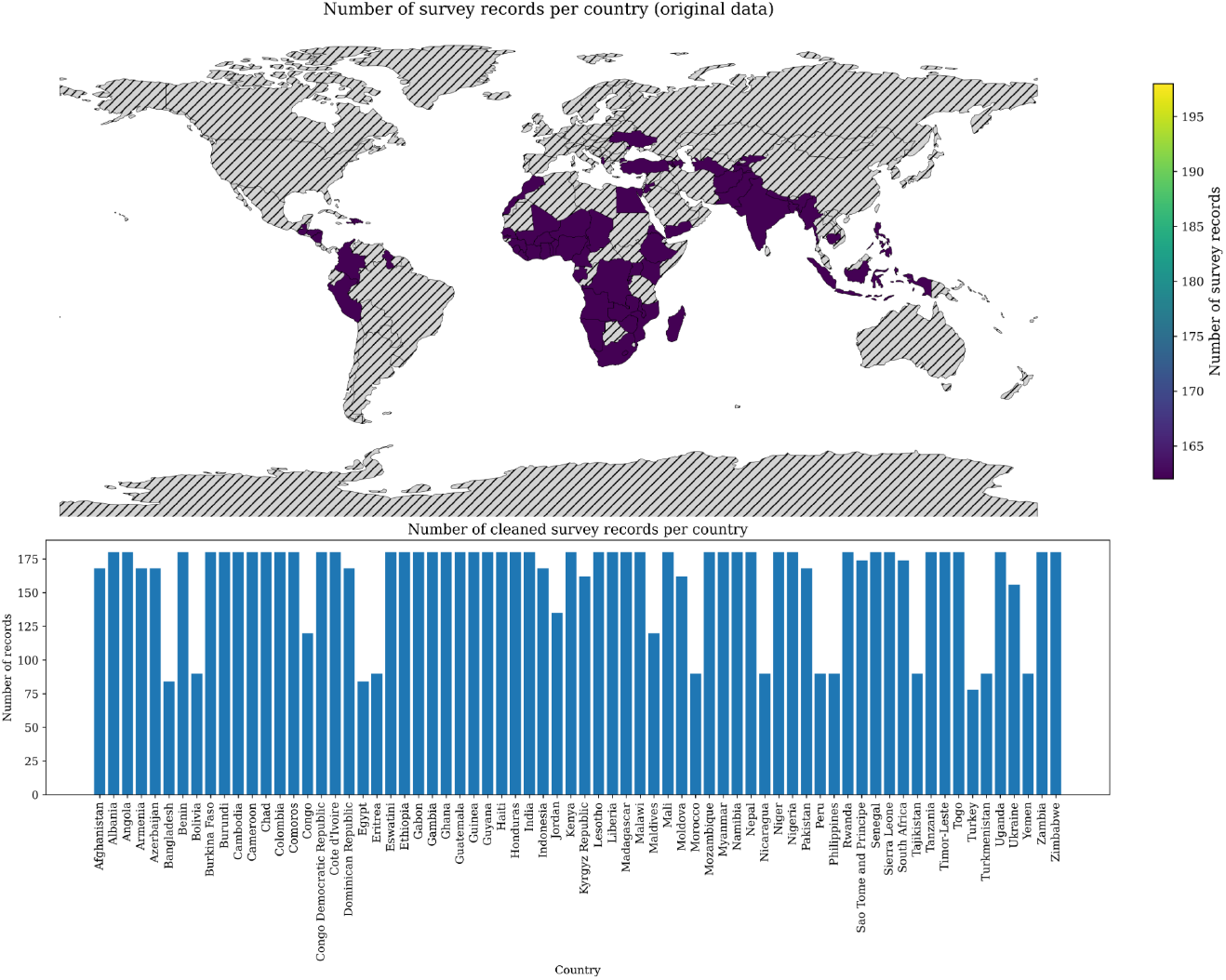
Country-level distribution of survey records. The upper panel shows the spatial distribution of available countries in the original survey compilation (uniform design of 175 records per country). The lower panel reports the number of cleaned records retained per country after validity filtering, illustrating heterogeneous data completeness across national surveys.

To isolate the relative contributions of demographic structure and attitudinal framing, we decomposed the modeling strategy into two complementary regimes. In Part I (Section 3.1), we trained per-country models using the full predictor set (demographics, survey question categories, year), thereby capturing how demographic composition and question framing jointly explain variation in violence-justification prevalence. In Part II (Section 3.2), we removed the survey question category predictors and trained per-country models using demographics only. This restricted specification provides a structural baseline that quantifies how much of the outcome can be explained without attitudinal question framing, enabling direct comparison between Part I and Part II.

#### 2.1.2 Country-level contextual integration (Part III)

For Part III (Section 3.3), to investigate whether the national patterns of violence acceptance learned from micro-level survey data are systematically related to broader societal structures, we integrated independent country-level indicators capturing cultural values, gender inequality, human development, and political regime quality (Table 3). These indicators were obtained from three globally established sources: Hofstede’s cultural dimensions [30], the United Nations Development Programme (UNDP) Human Development Reports [3], and the Varieties of Democracy (V-Dem) project [13] as disseminated via Our World in Data.^2^

**Table 3:** External macro–structural country-level indicators used in Part III, derived from Hofstede’s national cultural dimensions dataset, the UNDP Human Development Reports, and the V-Dem project as disseminated via OWID. **Abbreviations:** UNDP, United Nations Development Programme; V-Dem, Varieties of Democracy; OWID, Our World in Data.

| Category | Feature | Source | Interpretation |
| --- | --- | --- | --- |
| Cultural values | PDI | Hofstede | Power Distance Index (hierarchical inequality acceptance) |
|  | IDV | Hofstede | Individualism vs. collectivism |
|  | MAS | Hofstede | Masculinity vs. femininity orientation |
|  | UAI | Hofstede | Uncertainty Avoidance Index |
|  | LTO | Hofstede | Long-Term Orientation |
|  | IVR | Hofstede | Indulgence vs. restraint |
| Human development | HDE | UNDP | Human Development Index |
| Gender development | GDI | UNDP | Gender Development Index |
| Gender inequality | GII | UNDP | Gender Inequality Index |
| Democracy quality | Electoral-democracy | V-Dem/OWID | Electoral democracy quality |
|  | Liberal democracy | V-Dem/OWID | Liberal democracy quality |
| Women’s empowerment | Women political empowerment | V-Dem/OWID | Women’s political participation and power |

Hofstede’s cultural dimensions quantify persistent national value systems that structure social hierarchy, authority relations, and gender-role norms [31, 46, 54]. Power Distance (PDI) captures societal acceptance of hierarchical inequality [32]; Individualism (IDV) reflects the primacy of individual autonomy versus in-group conformity [47]; Masculinity (MAS) measures the cultural emphasis on dominance and gender differentiation [2]; Uncertainty Avoidance (UAI) indexes resistance to normative change [49]; Long-Term Orientation (LTO) reflects adherence to tradition versus adaptive future orientation [33]; and Indulgence (IVR) captures the degree of social regulation of personal autonomy [5]. Together, these dimensions operationalize latent cultural climates that may condition the societal acceptance of coercive control and violence-justifying beliefs [34].

The UNDP Human Development Reports provide a comprehensive, internationally comparable set of country-level composite indices that capture the broader socio-economic and institutional context of development [10, 44]. Among these indicators, the Human Development Index (HDI) integrates key dimensions of human well-being — education, health, and income — [60] while the Gender Development Index (GDI) and the Gender Inequality Index (GII) specifically quantify disparities in opportunity and outcomes between women and men [24]. These indices have been widely used in peer-reviewed research to link structural gender inequality and development with a range of health, social, and normative outcomes [67, 28].

For each country, we linked these macro-structural variables to the country-level violence acceptance index derived in Parts I–II, which summarizes the prevalence of endorsing violence-justifying attitudes across demographic strata. This integration enables a cross-scale analysis connecting individual-level normative beliefs to national cultural, developmental, and political conditions.

### 2.2 Country-resolved machine learning of violence-acceptance prevalence

In Parts I–II, violence-acceptance prevalence was formulated as a supervised regression problem and modeled separately for each country to capture country-specific predictive structure while avoiding pooling across heterogeneous survey contexts. Starting from the raw micro-level survey data, we applied a standardized preprocessing workflow—including type harmonization and exclusion of incomplete records—to produce a consistent modeling table comprising categorical predictors and survey year. Model development followed a fixed train/validation/test protocol (70%/15%/15%) and employed a unified preprocessing pipeline that one-hot encodes categorical variables and passes through numeric covariates, ensuring leakage-safe evaluation and reproducible estimation [9, 50].

For each country, we trained tree-based models that naturally handle sparse, high-dimensional one-hot representations: Random Forest regression, a single decision tree baseline, and gradient-boosted decision trees via XGBoost [6, 7, 12]. Hyperparameter optimization was performed using randomized cross-validated model selection [4, 59, 59]. For each country, hyperparameters were selected via *k*-fold cross-validation on the training subset only, followed by refitting on the combined training and validation data and final evaluation on a fully held-out test set to ensure unbiased performance estimation [65, 8]. The selected configuration was then retrained on the aggregated training–validation split and evaluated once on the independent test split.

Predictive performance was summarized using the coefficient of determination (*R*^2^) [71] as the primary metric, together with mean absolute error and root mean squared error [41]. The *R*^2^ quantifies the proportion of variance in the observed responses explained by the model and is defined in Eq. (1) [42, 45, 72].

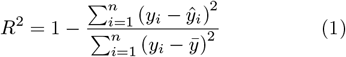

Root mean squared error (RMSE; Eq. (2)) computes the square root of the mean squared residuals and assigns greater weight to larger errors through quadratic penalization, thereby emphasizing the impact of large deviations [26, 23, 11]. To facilitate comparison across countries with different outcome scales, RMSE was normalized by the range of observed values within each country, yielding the normalized root mean squared error (NRMSE). While *R*^2^ captures relative variance explained but not the magnitude of prediction errors, it is commonly complemented with RMSE (and its normalized form, NRMSE) to assess absolute predictive accuracy and enable meaningful comparison between models with similar explained variance [42].

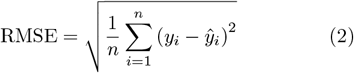

Part I used the full survey feature set available in the micro-level table, including the violence-acceptance item identifiers, whereas Part II restricted predictors to demographic variables only (gender, demographic question, demographic response, and survey year) while retaining the same country-specific modeling pipeline and evaluation protocol to isolate the demographic contribution to explainable variation in violence-acceptance prevalence [50, 12]. Results from these country-resolved analyses are presented in the subsequent Results sections, with Part I summarizing global performance patterns and model agreement (Section 3.1) and Part II examining demographic drivers of violence-justification attitudes (Section 3.2).

### 2.3 Country-level association analysis with macro-structural indicators

In Part III, we complemented the within-country supervised learning analyses (Parts I–II) with a country-level association framework designed to relate population-level acceptance of violence against women to broader societal conditions. For each country, the estimated prevalence of violence-justifying attitudes—aggregated across demographic strata—was linked to a set of external macro-structural indicators capturing gender inequality, human development, and political governance. Specifically, we integrated the most recent available values of the Gender Inequality Index, Gender Development Index, and Human Development Index, together with indices of electoral democracy, liberal democracy, and women’s political empowerment. All data sources were harmonized at the country level. A detailed description of the external data sources, indicators, and harmonization procedure is provided in sub-section 2.1.2.

Associations between national prevalence of violence-justifying attitudes and each external structural indicator were quantified using Spearman’s rank correlation, a nonparametric measure appropriate for monotonic relationships and robust to departures from normality [58, 57]. Statistical uncertainty was evaluated using two complementary inferential approaches. First, we conducted a Bayesian analysis of rank correlations based on a Fisher-*z* approximation with a weakly informative prior on the population correlation coefficient [19], yielding Bayes factors to quantify evidential strength and posterior probabilities characterizing the direction of association [35, 68]. Second, we applied a nonparametric bootstrap procedure to derive empirical 95% confidence intervals and to estimate the probability that the association is positive, providing a distribution-free assessment of robustness [18].

To support interpretation beyond global summary statistics, we examined country-specific contextual profiles using complementary visual summaries, as reported in subsection 3.3. These representations contrasted national levels of violence-justifying attitudes with multiple societal indicators, enabling comparative assessment across countries while preserving relative structure within the sample.

## 3 Results

### 3.1 Part I — Global performance patterns and model agreement

Across countries, predictive accuracy differed substantially when modeling the prevalence of violence-justifying attitudes from the full survey specification (demographics, attitudinal scenario item, and survey year; Figure 2). Test performance (*R*^2^) varied both by country and by model family (DT, RF, XGB; Figure 2A). Overall, XGBoost provided the strongest out-of-sample fit, achieving the highest test *R*^2^ in 65 of 70 countries (per-country summary table), motivating its use for the global summary in Figure 2B, which maps XGBoost test *R*^2^ across countries and reveals pronounced cross-national heterogeneity in predictability.

**Figure 2:**
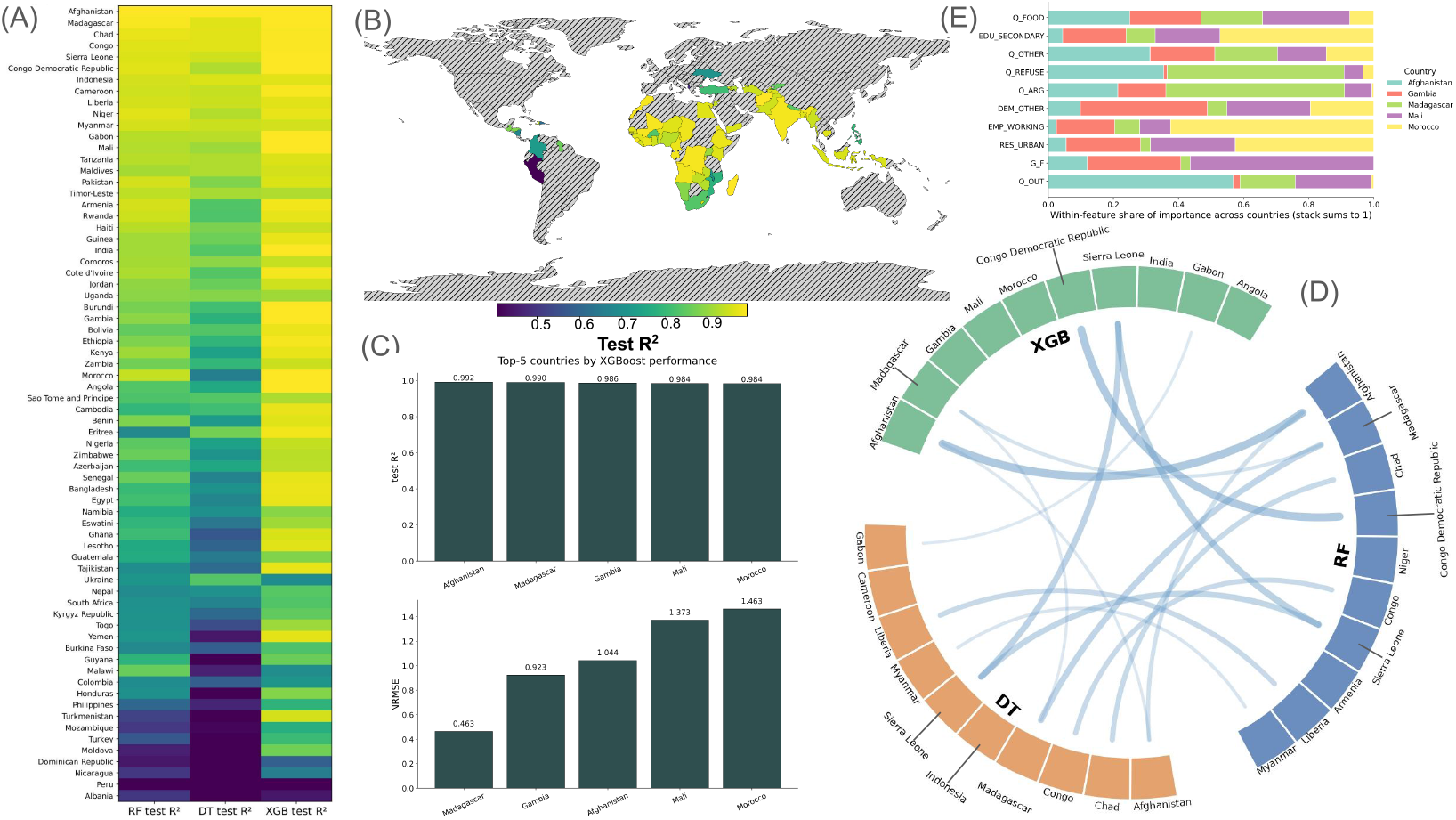
Part I: Cross-country predictive performance and feature attribution using the full survey specification. (A) Test-set *R*^2^ by country for Decision Tree (DT), Random Forest (RF), and gradient-boosted tree models (XGBoost; XGB), trained independently within each country using demographics, attitudinal scenario items, and survey year. (B) Global distribution of XGBoost test-set *R*^2^, illustrating cross-national heterogeneity in predictive performance under the full feature specification. (C) Top-five countries ranked by XGBoost test-set *R*^2^, with corresponding RMSE values on the held-out test set. (D) Cross-model overlap in top-ranked countries, linking countries that appear among the highest-performing sets for DT, RF, and XGBoost (XGB). (E) Predictor-family contribution profiles for the five highest-performing XGBoost (XGB) models. Within each country, raw one-hot feature importances were normalized and aggregated into interpretable feature families. Features are ordered by their average contribution across the five countries; stacked segments indicate the relative contribution of each country to a given feature family. **Abbreviations:** DT, Decision Tree; RF, Random Forest; XGBoost/XGB, Extreme Gradient Boosting; *R*^2^, coefficient of determination; RMSE, root mean squared error; *Q*_*FOOD*, justification if wife burns food; *Q*_*REFUSE*, justification if wife refuses sex; *Q*_*ARG*, justification if wife argues with husband; *Q*_*OUT*, justification if wife goes out without permission; *Q*_*OTHER*, endorsement for at least one reason; *EDU* _*SECONDARY*, secondary or higher education; *EMP* _*WORKING*, employed/working; *RES*_*URBAN*, urban residence; *RES*_*RURAL*, rural residence; *G*_*F*, female respondents.

Among the highest-performing settings under XG-Boost, Afghanistan (*R*^2^ = 0.982), Madagascar (*R*^2^ = 0.970), Georgia (*R*^2^ = 0.968), Mali (*R*^2^ = 0.961), and Morocco (*R*^2^ = 0.961) exhibited near-ceiling explained variance on the held-out test set (Figure 2C). Corresponding RMSE values highlight that high variance explained may coexist with non-negligible absolute error depending on the scale and dispersion of the outcome in each country.

Cross-model overlap in top-ranked countries is summarized in Figure 2D, where Sierra Leone appears among the top sets of all three estimators, whereas Afghanistan and the Democratic Republic of the Congo overlap between DT and RF. To examine which predictors drive the best-performing XGBoost models, Figure 2E aggregates one-hot encoded variables into interpretable feature families (attitudinal scenario items and demographic strata) and ranks them by average contribution across the five top-performing countries. In Afghanistan, Madagascar, Gambia, and Mali, attitudinal scenario items—particularly justification related to burning food (*Q*_*FOOD*), refusing sex (*Q*_*REFUSE*), arguing with the husband (*Q*_*ARG*), and going out without permission (*Q*_*OUT*)—dominate the highest ranks of feature importance. In contrast, Morocco shows a distinct profile in which demographic strata, especially educational attainment (*EDU* _*SECONDARY* and higher), employment status (*EMP* _*WORKING*), and place of residence (*RES*_*URBAN, RES*_*RURAL*), rank above most attitudinal items, while Mali exhibits an intermediate pattern combining both scenario-based and demographic contributions.

### 3.2 Part II — Demographic drivers of violence-justification attitudes

Under the demographics-only specification, predictive performance decreased across countries relative to the full survey models, yet remained non-negligible in a subset of cases (Figure 3). Among the highest-performing demographic-only XGBoost models, Ethiopia (*R*^2^ = 0.779), Niger (*R*^2^ = 0.755), Morocco (*R*^2^ = 0.674), Senegal (*R*^2^ = 0.602), and Benin (*R*^2^ = 0.580) achieved the largest explained variance on the held-out test set. Corresponding normalized RMSE values were 0.110, 0.128, 0.133, 0.143, and 0.159, respectively, confirming that demographic structure alone retains measurable predictive signal despite the exclusion of attitudinal scenario items.

**Figure 3:**
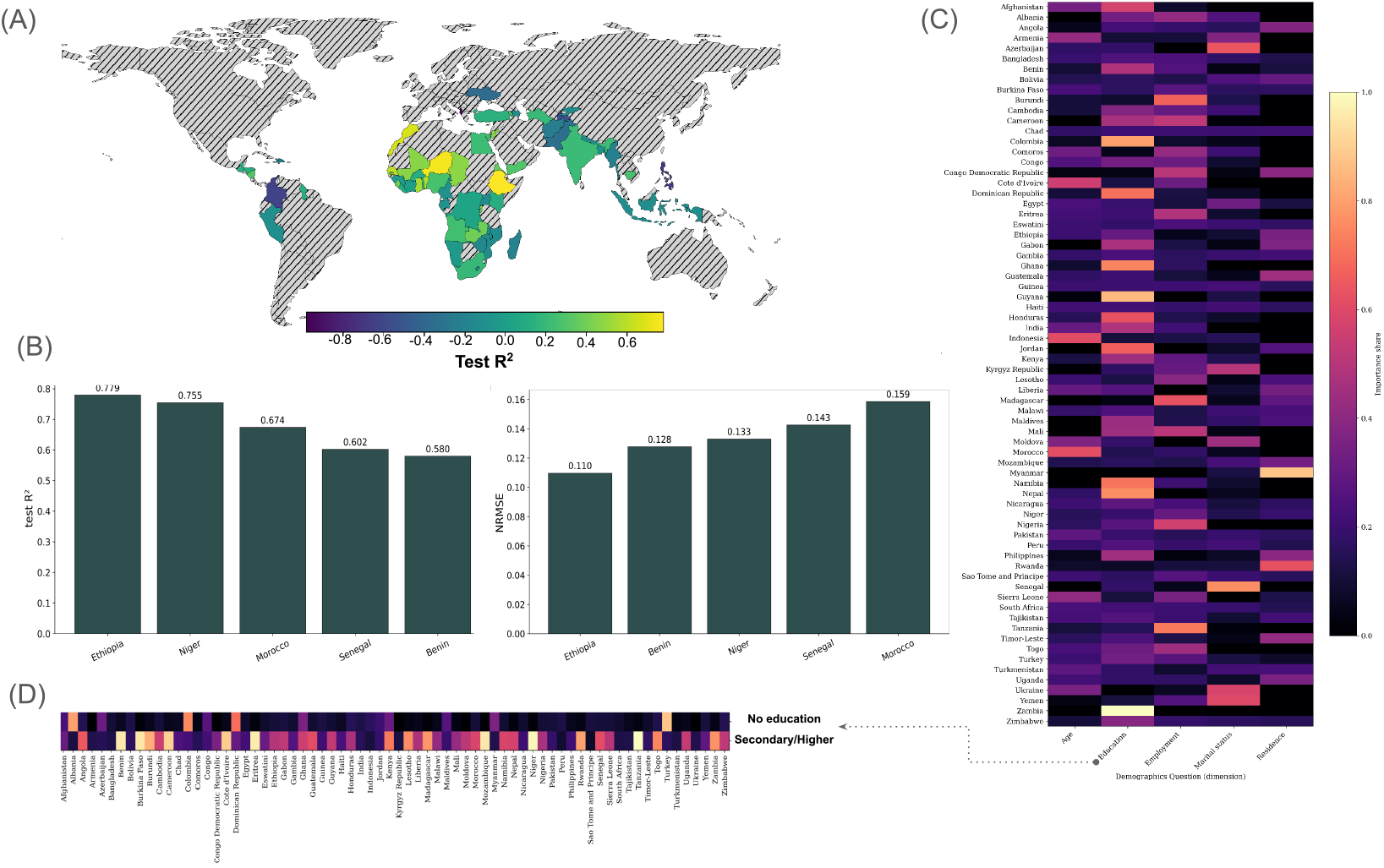
Part II — Demographics-only modeling of violence-justification prevalence. (A) Global distribution of XGBoost test performance (*R*^2^) under the demographics-only specification, in which attitudinal scenario items are excluded and predictions rely solely on demographic variables and survey year. Countries with insufficient data are shown in gray. (B) Test-set performance for the five highest-performing countries under the demographics-only model, reporting *R*^2^ (left) and normalized root mean squared error (NRMSE; right), highlighting substantial but reduced predictive accuracy compared with the full specification. (C) Heatmap of normalized XGBoost feature importance across countries, aggregated at the level of demographic feature families. Color intensity reflects the relative contribution of each demographic dimension to model predictions within each country. (D) Disaggregated view of education-related feature importance, showing the relative contribution of individual education response categories (from no education to higher education) across countries. Among these, the no education and secondary or higher education responses emerge as the two most influential isolated predictors, forming a consistent monotonic gradient in which lower educational attainment is associated with higher violence-justification prevalence, whereas higher education contributes most strongly to predictive differentiation.**Abbreviations:** XG-Boost, Extreme Gradient Boosting; *R*^2^, coefficient of determination; NRMSE, normalized root mean squared error.

The global distribution of demographics-only test *R*^2^ (Figure 3A) reveals substantial cross-national heterogeneity. While several countries retain moderate predictive accuracy based solely on demographic composition, many exhibit near-zero performance once attitudinal framing is removed, indicating that demographic structure alone is insufficient to explain violence-justification prevalence in those contexts.

Figure 3C–D further refines the role of education by resolving individual response categories within the demographics-only specification. A clear monotonic gradient is observed across countries, with responses corresponding to no or primary education showing stronger associations with higher prevalence of violence-justifying attitudes, whereas secondary and higher education categories contribute most strongly to predictive differentiation.

This pattern is corroborated by complementary descriptive analyses on the original survey data. Spearman rank correlations indicate a positive association between having no education and violence-justification prevalence (*ρ* = 0.248, *p* = 2.5 × 10^−42^), alongside a stronger negative association for secondary or higher education (*ρ* = −0.368, *p* = 5.8 × 10^−95^). Consistent with these rank-based associations, direct aggregation of responses across countries and demographic strata reveals a monotonic decrease in mean violence-justification prevalence with increasing educational attainment. Together, these observations indicate that education captures a structured and internally coherent demographic dimension, rather than an isolated category effect, and it represents the most robust demographic correlate of attitudes toward violence against women when attitudinal scenario framing is excluded.

### 3.3 Part III — Bayesian evidence for country-level hierarchy indicators

Figure 4A shows country-level associations between violence-justification prevalence and macro-structural indicators derived from the UNDP and the V-Dem/Our World in Data. Strong negative rank correlations are observed for the GDI, HDI, electoral democracy, and liberal democracy, indicating that countries with higher development levels and stronger democratic institutions tend to exhibit lower prevalence of violence-justifying attitudes. Among these indicators, GDI and HDI show the strongest associations, accompanied by substantial Bayesian evidence in favor of a non-zero relationship and comparatively narrow bootstrap confidence intervals, indicating robust and stable effects across countries.

**Figure 4:**
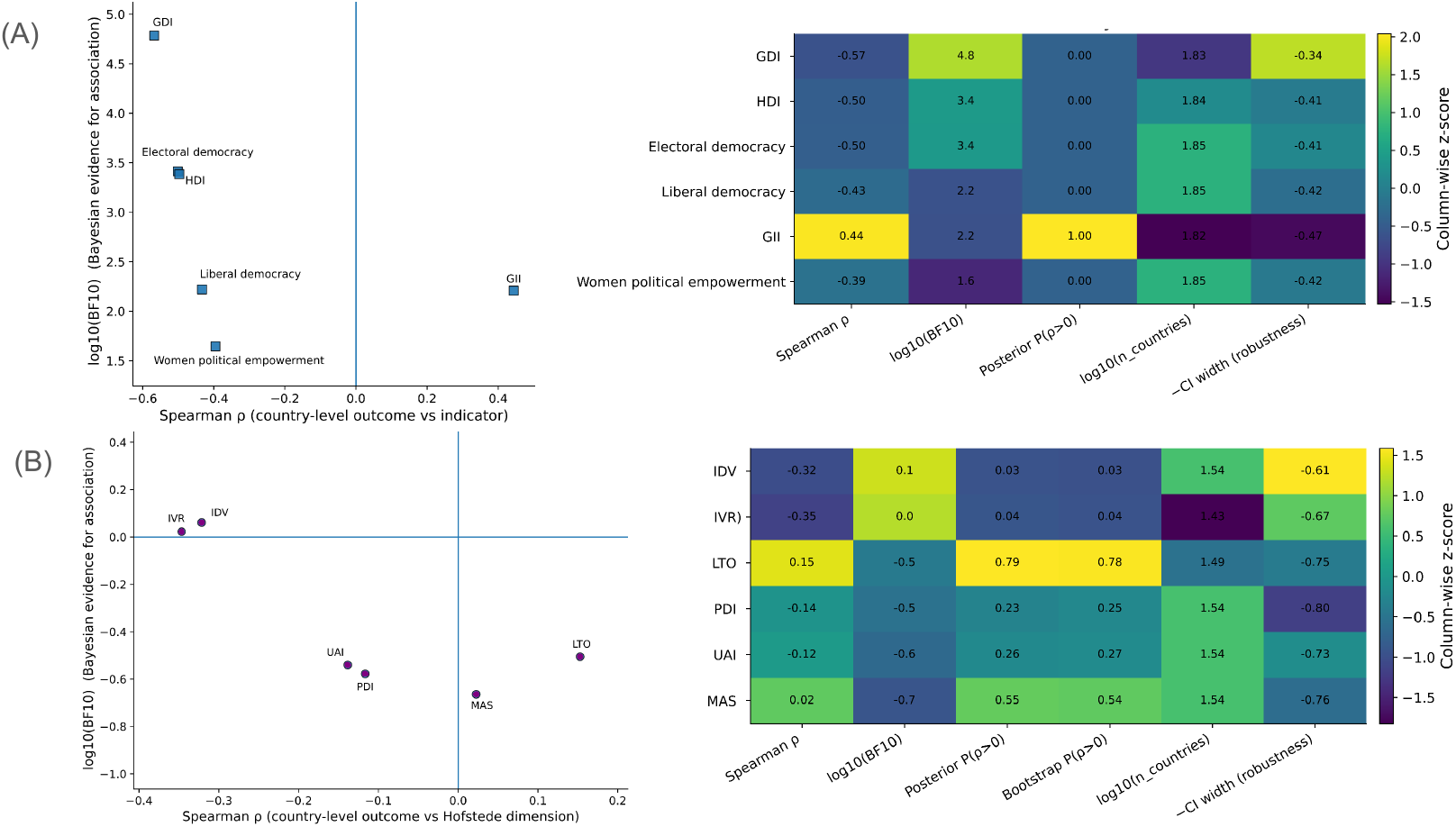
Country-level associations between violence-justification prevalence and macro-level indicators. (A) Associations between national prevalence of violence-justifying attitudes and structural indicators of development, gender inequality, and democratic quality. Points are positioned by Spearman rank correlation (*ρ*) and Bayesian evidence for association (log_10_(BF_10_)), with accompanying summaries of posterior probability, boot-strap robustness, sample size, and confidence interval width shown in the adjacent heatmap. (B) Corresponding association analysis for Hofstede’s cultural dimensions. Compared with structural indicators, cultural dimensions exhibit weaker correlations, lower Bayesian evidence, and reduced robustness, indicating more limited explanatory power for cross-national variation in violence-justifying attitudes. **Abbreviations:** *ρ*, Spearman rank correlation; BF_10_, Bayes factor in favor of association; HDI, Human Development Index; GDI, Gender Development Index; GII, Gender Inequality Index; PDI, Power Distance Index; IDV, Individualism; MAS, Masculinity; UAI, Uncertainty Avoidance Index; LTO, Long-Term Orientation; IVR, Indulgence versus Restraint.

In contrast, women’s political empowerment—also obtained from V-Dem—displays a weaker association with violence-justification prevalence, characterized by lower effect size and reduced Bayesian support. Overall, the pattern in Figure 4A highlights that broad structural conditions related to development, gender equality, and institutional quality are systematically linked to cross-national variation in attitudes toward violence against women.

Figure 4B presents the corresponding association analysis for Hofstede’s cultural dimensions. Compared with the structural indicators shown in Panel A, cultural value dimensions exhibit weaker and less consistent relationships with violence-justification prevalence. PDI and UAI show modest negative rank correlations, and MAS and LTO display small positive associations. IDV and IVR are weakly negatively associated with the outcome. Across all Hofstede dimensions, Bayesian evidence remains limited, and bootstrap confidence intervals are comparatively wide, indicating substantial uncertainty and reduced robustness relative to the macrostructural indicators.

To contextualize the countries exhibiting the highest predictive performance in Part I (Section 3.1), we examined their macro–structural profiles using the indicators identified in Part III as most strongly associated with violence-justification prevalence. As shown in Figure 4, development- and institution-based measures derived from the United Nations Development Programme (UNDP) and the Varieties of Democracy project (V-Dem/OWID) exhibit substantially stronger and more robust associations with the outcome than Hofstede’s cultural dimensions, motivating their use for country-level contextualization. Figure 5 summarizes this analysis for Afghanistan, Madagascar, Gambia, Malawi, and Morocco, which achieved the highest out-of-sample predictive accuracy under the full survey specification. Figure 5A shows standardized country-wise deviations of UNDP and V-Dem indicators, revealing pronounced structural heterogeneity among high-performing countries. Afghanistan is characterized by extreme disadvantage in gender development, human development, and democratic quality, whereas Morocco exhibits comparatively higher development and women’s political empowerment despite moderate gender inequality. Madagascar, Gambia, and Malawi occupy intermediate positions, reflecting distinct combinations of structural conditions. Figure 5B relates national violence-justification prevalence to each macro–structural indicator at the country level. Across indicators, higher prevalence is consistently associated with greater gender inequality and lower development and democratic quality. Countries such as Afghanistan and Morocco occupy extreme positions in these relationships, indicating that high predictive accuracy in Part I emerges in structurally polarized contexts rather than from uniformly similar national profiles.

**Figure 5:**
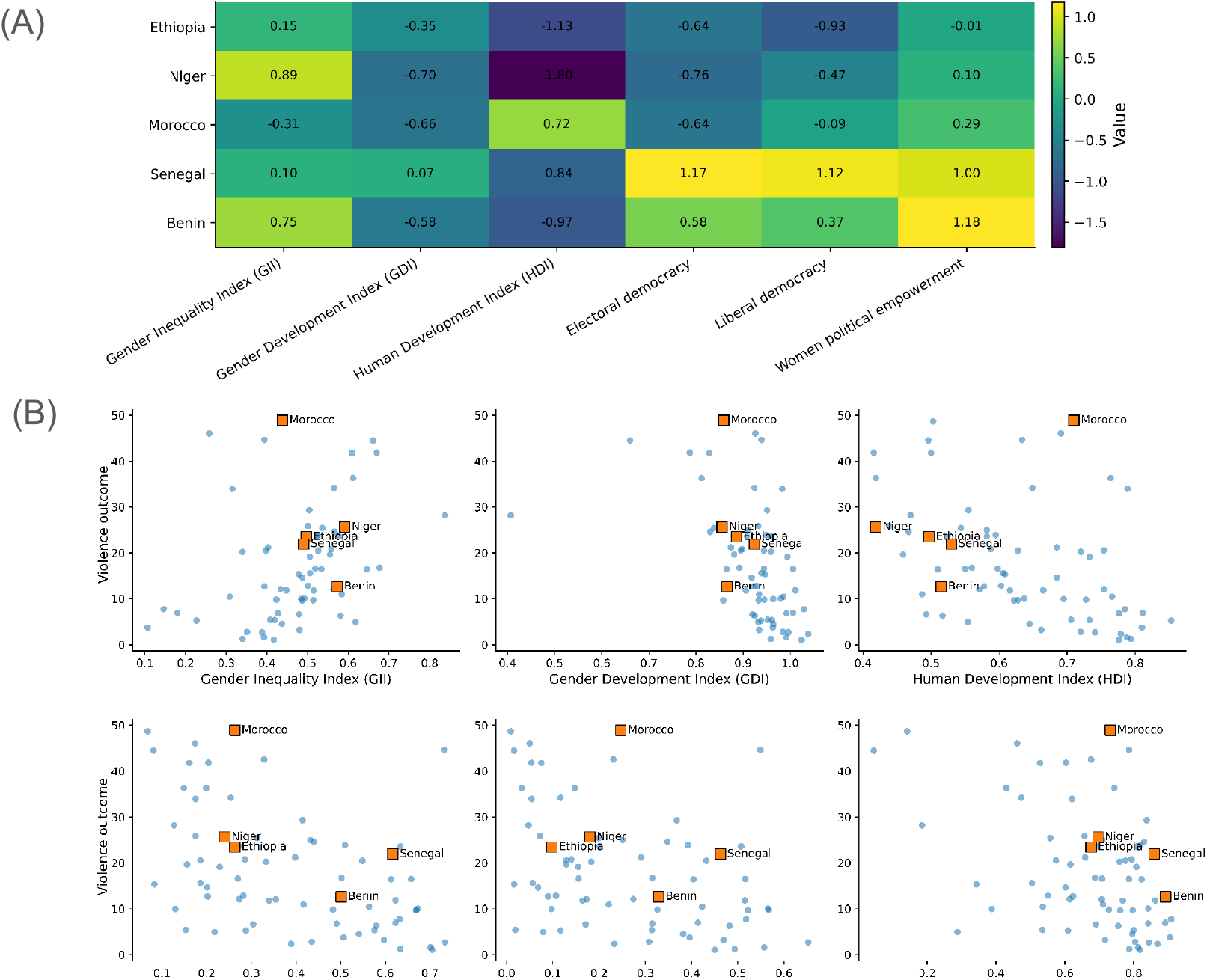
Macro–structural context of countries with highest predictive performance. Country-level association between violence-justification prevalence and macro–structural indicators for the five countries achieving the highest out-of-sample predictive accuracy in Part I (Afghanistan, Madagascar, Gambia, Malawi, and Morocco). (A) Standardized country-wise deviations (z-scores) of development, gender inequality, and democratic-quality indicators derived from the UNDP and the V-Dem/Our World in Data, highlighting pronounced heterogeneity in structural conditions across high-performing countries. (B) Country-level relationships between violence-justification prevalence and each macro–structural indicator, shown as scatter plots with highlighted high-performing countries. Across indicators, higher prevalence is associated with greater gender inequality and lower development and democratic quality. These country-specific patterns are consistent with the global association structure identified in Part III, where UNDP and V-Dem indicators exhibited stronger and more robust associations with violence-justification prevalence than cultural-value dimensions. **Abbreviations:** UNDP, United Nations Development Programme; V-Dem/Our World in Data, Varieties of Democracy project; HDI, Human Development Index; GDI, Gender Development Index; GII, Gender Inequality Index; BF_10_, Bayes factor in favor of association.

Figure 6 contextualizes the countries exhibiting the highest predictive performance under the demographics-only specification (Part II; Section 3.2) within the macro–structural dimensions identified in Part III. The analysis focuses on Ethiopia, Niger, Morocco, Senegal, and Benin, which achieved the highest out-of-sample accuracy when attitudinal scenario items were excluded.

**Figure 6:**
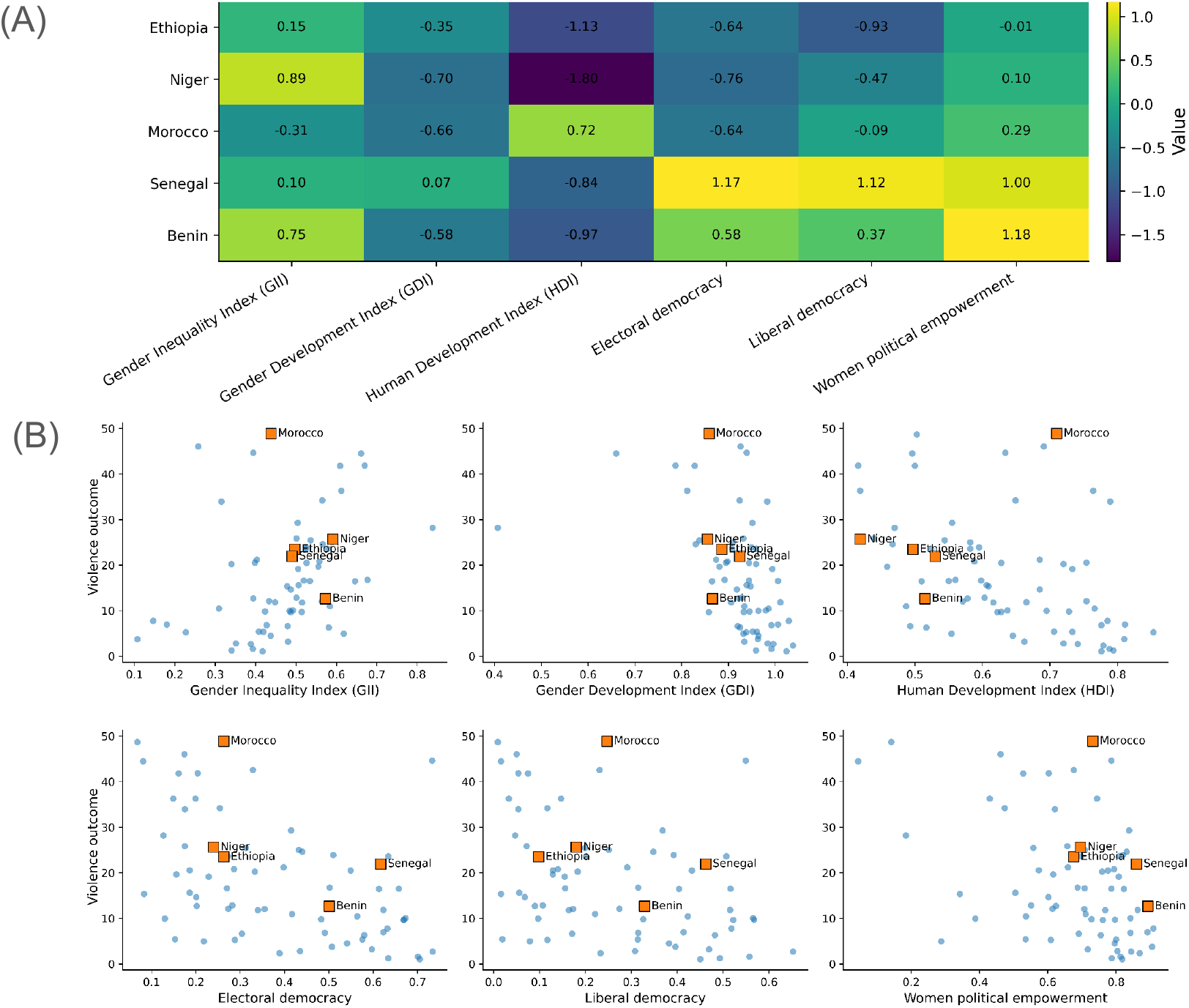
Macro–structural context of countries with highest predictive performance under the demographics-only specification. Country-level associations between violence-justification prevalence and macro–structural indicators for the five countries achieving the highest out-of-sample predictive accuracy in Part II (Ethiopia, Niger, Morocco, Senegal, and Benin). (A) Standardized country-wise deviations (z-scores) of development, gender inequality, and democratic-quality indicators derived from the UNDP and the V-Dem/Our World in Data, illustrating pronounced heterogeneity in structural conditions across high-performing countries. (B) Country-level relationships between violence-justification prevalence and each macro–structural indicator, shown as scatter plots with highlighted high-performing countries. As in Part I, higher prevalence is associated with greater gender inequality and lower development and democratic quality, indicating that demographic predictability remains embedded within sharply differentiated macro–structural environments. **Abbreviations:** UNDP, United Nations Development Programme; V-Dem/Our World in Data, Varieties of Democracy project; HDI, Human Development Index; GDI, Gender Development Index; GII, Gender Inequality Index; BF_10_, Bayes factor in favor of association.

Figure 6A shows standardized country-wise deviations of UNDP and V-Dem indicators, revealing marked heterogeneity among high-performing countries. Ethiopia and Niger exhibit a high GII, but a low GDI, HDI, and democratic quality. In contrast, Senegal and Benin exhibit comparatively higher electoral democracy, liberal democracy, and women’s political empowerment, while Morocco is characterized by higher human development and women’s political empowerment despite moderate gender inequality. These profiles indicate that strong demographic predictability arises across structurally diverse national contexts.

Figure 6B relates national violence-justification prevalence to each macro–structural indicator. Across indicators, higher prevalence is consistently associated with greater gender inequality and lower gender development, human development, and democratic quality. The highlighted high-performing countries occupy distinct but extreme positions along these gradients, confirming that demographic predictability in Part II is amplified in contexts with pronounced structural contrasts rather than uniform macro–structural conditions.

To assess whether high predictive performance in Parts I–II reflects shared macro–structural profiles, we applied hierarchical clustering to country-level indicators of development, gender inequality, democratic quality, and women’s political empowerment (Figure 7). Figure 7A shows that the highest-performing countries do not form a single homogeneous group but instead separate into distinct structural regimes, with Afghanistan strongly isolated, Morocco occupying a distinct branch with higher development and empowerment, and Benin, Malawi, and Senegal clustering at intermediate levels. Figure 7B extends this analysis to all countries, revealing that high-performing cases remain embedded within broader clusters defined by gradients of development and institutional quality rather than forming a unique group. Notably, Afghanistan appears as the least connected country in the similarity network, exhibiting substantially lower correlations with other countries (around 0.5), whereas the remaining high-performing countries show much stronger mutual similarity, with pairwise correlations typically exceeding 0.75.

**Figure 7:**
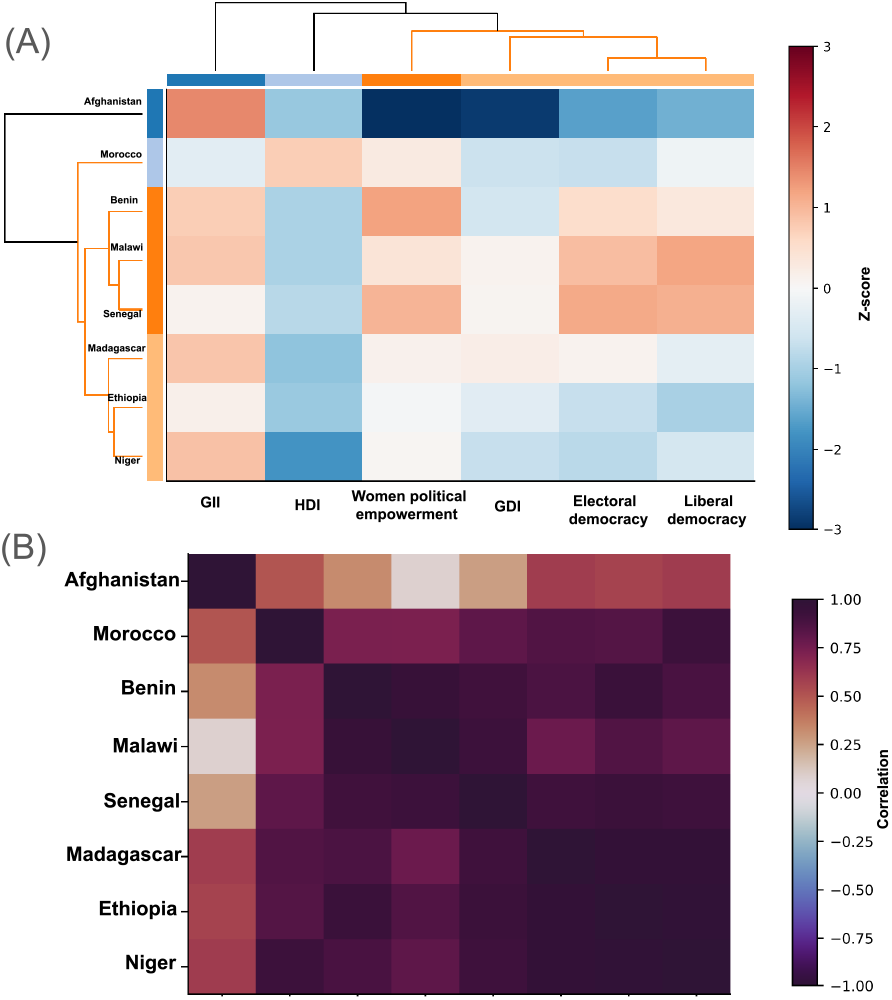
Country-level clustering by macro–structural indicators. Hierarchical clustering of countries based on development, gender inequality, democratic quality, and women’s political empowerment. (A) Clustering of countries with the highest predictive performance in Parts I–II, showing separation into distinct macro–structural profiles. (B) Clustering of all countries, where high-performing cases remain embedded within broader structural gradients rather than forming a distinct group. Afghanistan appears as the most structurally isolated country, whereas the remaining high-performing countries exhibit stronger mutual similarity. Heatmap values represent pairwise country similarity based on Spearman correlations across indicators. **Abbreviations:** HDI, Human Development Index; GDI, Gender Development Index; GII, Gender Inequality Index.

## 4 Discussion

### 4.1 Principal findings and conceptual contribution

This study reframes the normative acceptance of intimate-partner violence as a predictable population-level quantity, rather than solely an individual-level attitude or a correlate of victimization risk. Using DHS-derived harmonized country–year–subgroup prevalence estimates, we show that country-resolved supervised regression can explain a large fraction of held-out variation in violence-justification prevalence under the full survey specification (Part I; Figure 2). Crucially, the machine-learning framework is not used for individual risk prediction but as a diagnostic tool to quantify how much structure is present in normative endorsement patterns and how that structure differs across countries and survey designs. To support interpretability, model behavior is examined using feature-ranking analyses aggregated at the level of conceptually meaningful feature families, enabling direct assessment of the relative contributions of attitudinal framing and demographic structure (Figure 2E). The resulting predictability is strongly heterogeneous across countries (Figure 2B), and the highest-performing cases reach near-ceiling test *R*^2^ values (Figure 2C).

When attitudinal scenario items are removed (Part II), predictive accuracy declines globally (Figure 3A), yet remains substantial in a subset of countries (Figure 3B), indicating that demographic structure alone can retain measurable signal in some settings. Finally, in Part III, we find that country-level variation in violence-justification prevalence aligns more strongly with development-, gender-equality-, and institution-related indicators than with Hofstede-style cultural dimensions (Figure 4), and that countries with high predictive performance do not occupy a single uniform macro-structural regime (Figures 5–7).

### 4.2 Why predictability is high in some countries: scenario structure vs. demographic structure

A central empirical pattern is that, in several of the highest-performing countries under the full specification—Afghanistan, Madagascar, Georgia, Mali, and Morocco (Figure 2C)—feature-family attribution is dominated by attitudinal scenario identifiers (Figure 2E). This indicates that a sizable share of explainable variance is carried by the type of justification prompt (e.g., “burns food”, “refuses sex”, “argues”, “goes out”), rather than only by demographic structure. These justification items correspond to the attitudinal framing variables defined in Table 2, which capture scenario-specific normative conditions under which respondents report violence as justified. Substantively, this is consistent with a long DHS-based literature—introduced in the section 1—showing that endorsement levels differ systematically across specific wife-beating justification scenarios and that the scenario profile can vary markedly across national contexts and regions, even when using harmonized DHS instruments [64, 61, 38]. In these studies, scenario-conditional acceptance reflects structured normative distinctions about which situations are perceived as warranting coercion, and these distinctions differ across countries and subpopulations [29, 36]. Our results extend this descriptive and inferential evidence by demonstrating that scenario-conditioned structure can translate into strong out-of-sample predictability of country–subgroup prevalence when models learn from the full factorial survey design (Figure 2A–C).

Importantly, high predictability is not a single phenomenon with a single driver. Morocco serves as a counterpoint among the top performers (Figure 2C), exhibiting an attribution profile in which demographic strata (notably education, employment, and residence) rank above most scenario items (Figure 2E). This pattern aligns with prior cross-national evidence that socio-demographic gradients—especially education—are among the most robust correlates of accepting partner violence, with effects that can be substantial and context dependent [63, 17, 66]. In contrast, countries such as Afghanistan, Madagascar, and Mali show profiles in which scenario identifiers dominate (Figure 2E), suggesting that, in those settings, the prevalence surface is more strongly structured by which justification is being evaluated than by demographic segmentation alone.

This heterogeneity is further underscored in the demographics-only regime (Part II), where the highest-performing countries—Ethiopia, Niger, Morocco, Senegal, and Benin (Figure 3B)—retain non-trivial predictability even after scenario items are removed, indicating that demographic gradients alone can reconstruct part of the prevalence surface in specific contexts.

### 4.3 Education as the most robust demographic dimension under the baseline specification

Under the demographics-only specification, the clearest and most consistent signal is education (Figure 3C–D), forming a monotonic gradient in which lower educational attainment aligns with higher violence-justification prevalence while higher attainment contributes most strongly to predictive differentiation. This pattern closely aligns with a substantial body of evidence from Demographic and Health Surveys (DHS) and National Family Health Surveys (NFHS), which consistently identify education as one of the strongest and most stable correlates of attitudes toward—and experiences of—intimate partner violence. Across diverse contexts, women with no formal education are markedly more likely to justify or experience violence, whereas secondary or higher education is associated with substantially lower acceptance and risk [64, 61, 1]. For example, analyses of DHS data from Ghana show that women with higher education are significantly less likely to justify intimate partner violence compared with women without schooling, and that low education remains a robust predictor of increased violence risk even after accounting for partner behavior and household characteristics [17]. Longitudinal and multi-round NFHS analyses in India similarly document a persistent negative education gradient in both the justification and prevalence of physical and emotional violence over time, with acceptance and exposure highest among women with no formal education and lowest among those with secondary or higher schooling [1, 40]. Evidence from South Asia further indicates that education operates at the couple level: studies in Nepal, India, and Pakistan report that both women’s and partners’ educational attainment are inversely associated with domestic violence, with equally high-educated couples exhibiting the lowest likelihood of severe violence [40, 48]. Complementary work among adolescents and young adults in Bangladesh, India, and Nepal shows that lower educational attainment is associated with greater justification of wife beating, underscoring education’s role in shaping normative beliefs prior to marriage [61].

Importantly, the present findings extend this largely inferential literature by demonstrating that education retains predictive utility even when all attitudinal scenario framing is excluded. In Part II, education alone carries sufficient out-of-sample signal to reconstruct a substantial portion of the violence-justification prevalence surface in a subset of countries (Figure 3B), indicating that educational stratification captures a coherent structural dimension of normative acceptance rather than a collection of isolated associations. From this perspective, education emerges not only as a correlate of violence-related attitudes but also as a key organizing axis of their population-level structure.

### 4.4 Cross-scale embedding: institutions and development outperform compact culture indices

While education emerges as the dominant demographic axis where demographic stratification is present, Part II also reveals substantial cross-national heterogeneity in whether demographic structure alone explains violence-justification prevalence (Figure 3A). In many countries, removing attitudinal scenario framing collapses predictive performance toward zero, indicating that demographic segmentation alone is insufficient to explain prevalence variation in those contexts. This pattern is particularly evident in countries such as Afghanistan, which exhibits near-ceiling predictability under the full specification (Part I) but weak performance under the demographics-only model (Part II).

As shown in the standardized macro–structural heatmaps in Figure 5A, Afghanistan occupies an extreme position relative to the full country sample, with substantially worse-than-average scores on gender inequality, human and gender development, democratic quality, and women’s political empowerment. Here, standardized z-scores quantify each country’s deviation from the sample mean on a given macro– structural indicator, providing a positional—rather than causal—context for interpreting predictive performance. In such structurally extreme settings, endorsement of violence-justifying norms appears relatively homogeneous across demographic strata once scenario framing is ignored, consistent with norms that are broadly shared rather than sharply stratified. Correspondingly, Afghanistan appears as a structurally isolated case in the clustering analysis (Figure 7), exhibiting low similarity to other high-performing countries in macro–structural space.

By contrast, countries such as Morocco retain substantial predictive accuracy under both the full and demographics-only specifications. As illustrated in Figs. 5 and 6, Morocco occupies a less extreme macro– structural position relative to the sample, with human and gender development and women’s political empowerment closer to—or modestly above—the sample mean, and markedly higher than in structurally isolated cases such as Afghanistan. This relative positioning is visible both in the z-score heatmaps and in the country-level association plots, where Morocco lies toward the higher end of the development and institutional gradients within the sample. In this context, demographic gradients—particularly education—remain informative even in the absence of attitudinal framing, indicating greater heterogeneity in normative acceptance across demographic groups. Consistent with this interpretation, Morocco clusters more closely with other structurally intermediate or higher-positioned countries in the similarity analysis (Figure 7), despite differing levels of violence-justification prevalence.

Taken together, these patterns indicate that whether demographic structure alone explains violence-justification prevalence depends on broader macro– structural positioning within the global sample, including relative levels of educational inequality, gender inequality, and institutional development. Finally, in Part III, we find that country-level variation in violence-justification prevalence aligns more strongly with development-, gender-equality-, and institution-related indicators than with Hofstede-style cultural dimensions (Figure 4). Standardized z-score profiles and clustering analyses further demonstrate that countries with high predictive performance occupy heterogeneous positions in macro–structural space rather than forming a single uniform regime (Figs. 5–7). To the best of our knowledge, prior DHS-mand NFHS-based studies have not explicitly examined this distinction using out-of-sample predictability, nor tested how the presence or absence of demographic structure itself varies systematically across countries. Future work could formally test these relationships by modeling demographic-only predictability as a function of macro–structural indicators, and by assessing whether structural extremity or embeddedness in development–institution space moderates the emergence of demographic gradients in normative acceptance.

### 4.5 Limitations

Several limitations qualify interpretation. First, the outcome is a prevalence estimate rather than individual-level responses, so results should not be read as individual risk prediction. Second, the number of observations per country is modest (median near the full factorial design but reduced heterogeneously; Figure 1), which increases variance in country-specific generalization estimates and may amplify sensitivity to how survey years and strata are represented. Third, scenario variables are measurement prompts: their predictive dominance in Part I reflects scenario-specific endorsement structure but does not itself identify mechanisms or direct policy levers. Fourth, in Part III, country-level associations are correlational; while robust rank-based and Bayesian evidence summaries are used, causal claims would require stronger identification strategies, including quasi-experimental designs or longitudinal within-country analyses.

### 4.6 Implications and future directions

Despite the limitations noted above, the findings point to several actionable implications with direct relevance for violence-prevention policy and for the design of norm-focused interventions. Methodologically, the country-resolved modeling framework introduced here provides a principled way to quantify how much of the population-level prevalence of violence-justifying attitudes is structurally explainable from survey composition alone and to decompose this explainability into scenario-driven versus demographic-driven components (Figs. 2–3). This distinction is substantively meaningful rather than purely technical: it differentiates contexts in which normative endorsement is primarily structured by which forms of violence are conditionally justified from those in which it is structured by who is more likely to endorse violence across demographic strata.

Substantively, the consistent role of education under the demographics-only specification (Part II; Figure 3C–D) reinforces its central importance as a structural correlate of violence-justifying norms. A large body of prior research has established that higher educational attainment is associated with lower acceptance of intimate-partner violence across diverse settings, even after accounting for wealth, residence, and employment [64, 61]. Our results extend this literature by demonstrating that education is not only statistically associated with attitudes but in some countries carries sufficient out-of-sample predictive signal to reconstruct a substantial portion of the prevalence surface in the absence of attitudinal framing. This finding supports existing evidence that sustained investments in female education and school participation can contribute to long-term normative change by reshaping beliefs about gender roles, authority, and the legitimacy of coercion [27, 43, 25].

At the same time, the dominance of scenario framing in several high-performing countries under the full specification (Part I; Figure 2E) indicates that demographic targeting alone is unlikely to be sufficient in many contexts. Where endorsement varies sharply across justification conditions (e.g., burns food versus refuses sex), effective prevention may require interventions that explicitly confront the moral narratives and conditional norms that render specific acts of violence socially acceptable. This interpretation is consistent with qualitative and mixed-methods evidence showing that violence-prevention programs are more effective when they address context-specific justifications and everyday moral reasoning, rather than framing violence solely as a generalized or abstract harm [37, 20].

The cross-scale associations identified in Part III further underscore the importance of institutional and developmental context. The strong and robust relationships between violence-justification prevalence and indicators of gender development, human development, and democratic quality (Figure 4) suggest that normative acceptance is embedded within broader opportunity structures, legal protections, and enforcement environments. Prior work has emphasized that legal reforms, women’s political representation, and institutional accountability play a critical role in shifting social norms around violence, particularly when combined with educational and community-based interventions [28, 39]. Our findings suggest that these macro– structural conditions may also shape how internally co-herent—and therefore how predictable—normative endorsement patterns are within countries.

From a practical perspective, the modeling framework proposed here could be used as a diagnostic tool to identify countries or subpopulations in which violence-justifying attitudes exhibit strong internal structure and may therefore be more responsive to targeted normative interventions. Countries characterized by high scenario-driven predictability may benefit most from campaigns that directly challenge conditional justifications of violence, whereas contexts with strong demographic gradients may require policies focused on educational access, economic participation, and the reduction of urban–rural disparities. Importantly, high predictability should not be interpreted as inevitability; rather, it signals the presence of a coherent normative structure that may be especially vulnerable to well-designed, context-specific disruption.

Future work can build on the present framework in several directions that directly address current limitations. First, explicitly modeling interactions between scenario framing and demographic strata would allow identification of the most normatively sensitive combinations, moving beyond marginal effects. Second, retaining complete survey years within countries would facilitate a formal evaluation of temporal generalization and the enduring stability of normative structures over time. Third, integrating policy and legal indicators—such as the enforcement of domestic violence legislation, access to survivor services, and compulsory education policies—would help link normative acceptance more directly to institutional mechanisms of protection and prevention. Together, these extensions would strengthen causal interpretation and enhance the framework’s utility for policy-relevant analysis.

## Acknowledgements

The author also acknowledges Zonta District 14 for its sustained engagement in initiatives addressing violence against women, which provided thematic motivation for this work. These contributions did not influence the study design, analysis, or interpretation of results. The author thanks Marius Oechner for suggesting the inclusion of Hofstede’s cultural indicators as contextual data.

## Declaration of generative AI and AI-assisted technologies in the writing process

During the preparation of this work, the author used ChatGPT to improve readability. After using this tool, the author reviewed and edited the content as needed and takes full responsibility for the content of the published article.

## Funding

This research received no external funding.

## Data availability

All data used in this study are derived from publicly available sources. The harmonized survey dataset analyzed in this work is available via the Demographic and Health Surveys Program and its public aggregations. External country-level indicators were obtained from publicly accessible databases, including the United Nations Development Programme and the Varieties of Democracy project. Detailed data sources and links are provided in the Materials and Methods section.

## Code availability

The code used for data preprocessing, modeling, and analysis is available from the author upon reasonable request.

## Author contribution

The author solely conceived the study, performed the analysis, interpreted the results, and wrote the manuscript.

## Footnotes

1 https://www.kaggle.com/datasets/whenamancodes/violence-against-women-girls

2 Hofstede cultural dimensions (Kaggle mirror used in this study): https://www.kaggle.com/datasets/seydakaba/hofstede-cultural-dimensions-by-country. UNDP Human Development Reports downloads: https://hdr.undp.org/data-center/documentation-and-downloads. Our World in Data Grapher endpoints for V-Dem indicators used here: https://ourworldindata.org/grapher/electoral-democracy-index.csv, https://ourworldindata.org/grapher/liberal-democracy-index.csv, https://ourworldindata.org/grapher/women-political-empowerment-index.csv.

## Notes

### Competing Interest Statement

The authors have declared no competing interest.

### Funding Statement

This study did not receive any funding

### Author Declarations

The study used only openly available human data that were originally obtained from the Demographic and Health Surveys (DHS) Program and publicly accessible aggregated compilations of DHS data.

